# Genomic Classification of Acute Lymphoblastic Leukemia Using AI: Towards Personalized Medicine

**DOI:** 10.1101/2025.09.20.25336225

**Authors:** Alireza Rahi, Mohammad Hossein Shafiabadi

## Abstract

Acute lymphoblastic leukemia is a highly heterogeneous hematologic malignancy that poses significant challenges for clinicians in terms of early detection and accurate subtype classification.

Misclassification or delayed recognition of molecular subtypes can lead to suboptimal treatment decisions and negatively impact patient outcomes. Traditional diagnostic methods, while effective, often lack the sensitivity to fully capture the underlying molecular diversity of acute lymphoblastic leukemia.

In this study, we applied an artificial intelligence–driven diagnostic framework to analyze gene expression profiles of patients with acute lymphoblastic leukemia. By leveraging advanced computational techniques, the model was able to identify distinct molecular signatures and stratify patients according to their genomic subtypes.

The findings demonstrate that AI-assisted genomic classification has the potential to enhance diagnostic precision and enable earlier identification of clinically relevant subgroups. This approach may support more tailored therapeutic strategies and represents an important step toward the integration of precision medicine in the management of acute lymphoblastic leukemia.

## Introduction

Acute lymphoblastic leukemia is one of the most common pediatric hematologic malignancies worldwide, characterized by high heterogeneity at the molecular and cellular levels [2], [7], [8]. Accurate and early classification of its molecular subtypes remains a significant challenge for clinicians, as misclassification can lead to suboptimal treatment strategies and adversely affect patient outcomes [9], [10]. Traditional diagnostic methods, including flow cytometry, cytogenetics, and singlegene testing, provide valuable information but often fail to fully capture the complex genomic landscape of acute lymphoblastic leukemia [11].

Recent advances in high-throughput gene expression profiling have enabled the collection of large-scale genomic datasets, which offer unprecedented opportunities for understanding disease heterogeneity and discovering clinically relevant biomarkers [1], [7]–[11]. Despite these advances, integrating such extensive data into reliable and interpretable diagnostic tools remains a formidable task, especially when accounting for batch effects, imbalanced representation of subtypes, and missing values across multiple studies.

Artificial intelligence techniques, particularly deep learning models such as convolutional neural networks, long short-term memory networks, and fully connected dense architectures, have demonstrated remarkable performance in pattern recognition tasks within biomedical datasets [2]–[6]. These models can capture complex, non-linear relationships in gene expression profiles and have been successfully applied to various cancers, including acute myeloid leukemia, skin cancer, and other malignancies [2]–[5]. However, the application of such models to acute lymphoblastic leukemia subtyping has been limited, partly due to the scarcity of balanced and high-quality datasets.

To address these challenges, we constructed a global reference balanced gene expression dataset for acute lymphoblastic leukemia subtypes, integrating 13 publicly available GEO microarray datasets and harmonizing 258 samples across nine molecular subtypes and normal controls [1]. Leveraging this dataset, we implemented an ensemble diagnostic framework combining convolutional neural networks, long shortterm memory networks, and dense neural networks to extract complementary genomic features. A gradient boosting classifier was subsequently trained as a meta-learner to integrate these features and provide accurate subtype predictions.

The proposed approach demonstrates the potential of artificial intelligence–driven], genomic classification to enhance diagnostic precision, facilitate early detection of clinically relevant subgroups, and support personalized therapeutic strategies. By combining advanced computational modeling with high-quality gene expression data, this study provides a critical step toward the integration of precision medicine into routine clinical practice for acute lymphoblastic leukemia.

## Related Work

Machine learning and deep learning approaches have increasingly been applied to the classification and prognosis of hematologic malignancies. Convolutional neural networks have demonstrated high performance in pattern recognition tasks across multiple biomedical datasets, including imaging and gene expression data [2], [4]. In dermatology, CNNs have achieved dermatologist-level accuracy in classifying skin cancers, demonstrating the ability of deep architectures to learn complex, non-linear features from highdimensional input data [2].

Long short-term memory networks have been successfully employed for modeling sequential and temporal relationships in biomedical signals and genomic data [3], [6]. For instance, LSTM architectures have been applied to gene expression profiles to capture dependencies among genes and improve subtype classification in leukemia [6]. Fully connected dense neural networks, although simpler in architecture, have also been effective for integrating complementary features and achieving high classification accuracy when combined in ensemble frameworks [4], [5].

Several studies have highlighted the challenges of applying deep learning models to leukemia classification, particularly due to limited sample sizes, batch effects, and imbalanced representation of subtypes [7]– [11]. Recent efforts to construct harmonized and balanced datasets have mitigated some of these issues, allowing models to generalize more effectively across molecular subtypes [1], [7]–[11]. The integration of multiple model predictions through ensemble learning and meta-learner strategies has further enhanced the robustness and accuracy of subtype classification [2]–[6].

Our approach builds upon these prior studies by leveraging a high-quality, balanced gene differences expression dataset and combining multiple deep learning architectures with a gradient boosting meta-learner. This strategy captures complementary genomic patterns and provides a reliable framework for accurate classification of acute lymphoblastic leukemia subtypes, which is essential for guiding personalized therapeutic interventions.

## Materials and Methods

### Dataset Construction

A global reference balanced gene expression dataset for acute lymphoblastic leukemia subtypes was curated by integrating 13 publicly available GEO microarray datasets, resulting in a harmonized collection of 258 samples across nine molecular subtypes and normal controls [1], [7]–[11]. The dataset contains 54,675 genes and was carefully balanced to ensure equitable representation of each subtype, thereby minimizing potential bias in downstream machine learning analyses.

### Data Preprocessing

Raw gene expression data were log-transformed and standardized using z-score normalization to reduce scale among genes [1]. Missing values were imputed using median values, and features with excessive missing values were removed. After cleaning, the dataset underwent further standardization using a StandardScaler to ensure zero mean and unit variance across all features [1], [3].

### Feature Selection

To reduce dimensionality and enhance model interpretability, the top 5000 genes were selected using the Analysis of Variance (ANOVA) F-test (SelectKBest) with respect to subtype labels [1]. This approach prioritizes genes with the highest discriminatory power for acute lymphoblastic leukemia subtypes, while maintaining sufficient genomic coverage.

### Deep Learning Architectures

Three complementary deep learning models were implemented: convolutional neural networks, long short-term memory networks, and fully connected dense networks [2]–[6].

1. **Convolutional Neural Network (CNN)**: Captures local patterns and dependencies in gene expression profiles using one-dimensional convolutional layers, followed by batch normalization, max-pooling, and dropout to prevent overfitting [2], [4].
2. **Long Short-Term Memory Network (LSTM)**: Models sequential dependencies among genes, capturing long-range correlations that may be indicative of subtype-specific expression patterns [3], [6].
3. **Dense Neural Network**: Integrates complementary information via fully connected layers and acts as a baseline model for ensemble learning [4], [5].

Each model was compiled using categorical cross-entropy loss and the Adam optimizer, and class weights were applied to account for imbalances among subtypes [1].

### Data Augmentation and Generators

To increase the effective sample size and improve model generalization, mild data augmentation was applied. Gaussian noise and small scaling perturbations were added to gene expression vectors [1]. A generator function yielded batches of augmented data for model training, ensuring continuous shuffling and efficient memory usage.

### Ensemble Learning and Meta-Learner

Predictions from the three deep learning models were concatenated to form metafeatures. A gradient boosting classifier was trained on these meta-features to act as a meta-learner, combining complementary information and improving overall subtype classification accuracy [2]–[6].

### Model Evaluation

The models were evaluated using multiple metrics, including accuracy, precision, recall, F1-score, and area under the ROC curve (AUC). Confusion matrices and ROC curves were generated to visualize classification performance across subtypes [1], [2], [4].

## Results

### Classification Performance

The ensemble model combining convolutional neural networks, long shortterm memory networks, and dense networks with a gradient boosting meta-learner achieved strong performance across the acute lymphoblastic leukemia subtypes. Table 1 summarizes the key metrics for each subtype.

**Table 1.** Classification Metrics per Subtype

| Subtype | Precision | Recall | F1-score | Support |
| --- | --- | --- | --- | --- |
| Acute Lymphoblastic Leukemia TEL-AML1 | 0.80 | 1.00 | 0.89 | 4 |
| Acute Lymphoblastic Leukemia Hyperdiploid | 0.80 | 0.67 | 0.73 | 6 |
| T-cell Acute Lymphoblastic Leukemia | 0.60 | 1.00 | 0.75 | 3 |
| Acute Lymphoblastic Leukemia E2A-PBX1 | 1.00 | 1.00 | 1.00 | 2 |
| Acute Lymphoblastic Leukemia Other | 0.80 | 0.80 | 0.80 | 5 |
| Acute Lymphoblastic Leukemia Ph-positive | 0.80 | 1.00 | 0.89 | 4 |
| Acute Lymphoblastic Leukemia MLL-rearranged | 1.00 | 0.25 | 0.40 | 4 |

#### Overall metrics

- Accuracy: 78.57%
- Mean ROC-AUC: 0.9685

#### Key Observations

- **ROC-AUC 0.9685** indicates excellent ability to discriminate positive versus negative cases across all subtypes. The ROC curve is close to the top-left corner, showing the model’s strong sensitivity and specificity.
- **High precision:** Most subtypes have precision above 80%, demonstrating reliable positive predictions.
- **Outstanding performance:**
- Acute Lymphoblastic Leukemia E2A-PBX1 achieved 100% across all metrics (Precision, Recall, F1-score).
- TEL-AML1 and Ph-positive subtypes reached 100% recall, indicating all positive cases were correctly identified. **Areas for improvement:**
- MLL-rearranged subtype shows only 25% recall, suggesting additional model optimization or more data may be necessary.
- Hyperdiploid subtype has 67% recall, indicating moderate sensitivity and room for enhancement

### Interpretation

The ensemble strategy effectively combines complementary predictions from CNN, LSTM, and Dense networks, resulting in high overall discrimination as reflected by the ROC-AUC score [2]–[6]. While most subtypes are classified with high accuracy, certain rarer or more heterogeneous subtypes (e.g., MLL-rearranged) remain challenging. Future work could include targeted augmentation strategies, feature selection refinement, or additional molecular markers to further improve recall for these subtypes [1], [7]–[11].

### CNN Model Analysis

The CNN model shows unstable training behavior.

- Training accuracy increases gradually but remains low (around 40%).
- Validation accuracy plateaus very early (around 20%), showing almost no improvement after a few epochs.
- The training loss decreases steadily, while the validation loss decreases slightly but remains significantly higher. [2]–[6]

This indicates **underfitting and poor generalization**, suggesting that the CNN architecture is not well-suited for this dataset or requires tuning (e.g., deeper layers, regularization, or more data) [2]–[6].

### Dense Model

The Dense (fully connected) model demonstrates much stronger performance.

- Both training and validation accuracy increase rapidly and stabilize around **70%**.
- Training and validation curves follow a similar trend, indicating a good fit without severe overfitting.
- Loss values (both training and validation) decrease consistently over epochs [2]–[6].

This suggests that the **Dense model is more effective** than CNN or LSTM for this task, capturing the underlying patterns in the data more successfully [2]–[6].

### LSTM Model

The LSTM model behaves similarly to the CNN model, with limited improvement.

- Training accuracy rises slowly but stays below **40%**.
- Validation accuracy plateaus around **20%**, with almost no improvement.
- Training loss decreases steadily, while validation loss remains nearly flat [2]–[6]. This indicates that **the LSTM is not effective** for this dataset in its current configuration. It suffers from underfitting, possibly due to insufficient sequence-related patterns in the data or the need for hyperparameter tuning [2]–[6].

### Ensemble Meta-Learner Performance

Despite the moderate individual performance of the base learners (CNN, Dense, and LSTM), the ensemble metalearner successfully combined their predictions, achieving a final accuracy of 78.57% and an excellent mean ROC-AUC score of 0.9685 [1], [2]–[6]. This clearly demonstrates the strength of ensemble methods in leveraging complementary strengths of multiple models, even if some base learners perform suboptimally, to build a more robust and accurate final predictor [1], [2]–[6].

### Confusion Matrix Interpretation

The confusion matrix (Figure 4) demonstrates the classification performance of the ensemble model (CNN + LSTM + Dense + Gradient Boosting) across **seven major ALL subtypes**. The model achieves strong performance in correctly identifying **ALL_TEL_AML1, T_ALL, ALL_E2A_PBX1, ALL_other, and ALL_Ph_positive**, where the diagonal dominance indicates effective subtype discrimination. For instance, **ALL_TEL_AML1** was predicted with perfect recall (4/4 correct), while **ALL_E2A_PBX1** achieved both **100% precision and recall**.

However, misclassifications were observed in **ALL_Hyperdiploid** (1 case predicted as ALL_other) and particularly in **ALL_MLL_rearranged**, where the model struggled, with only **1 correct prediction out of 4**. These errors suggest challenges in distinguishing genetically complex or overlapping subtypes, which aligns with findings in previous leukemia transcriptomic studies [7], [8], [9].

### ROC Curve and AUC Performance

The ROC curve (Figure 2) highlights the discriminative power of the model across subtypes. The **mean ROC-AUC of 0.9685** confirms the robustness of the ensemble method. Several subtypes, such as **T_ALL, ALL_E2A_PBX1, and ALL_Ph_positive**, reached a **perfect AUC of 1.000**, while **ALL_TEL_AML1 and ALL_other** achieved **AUC values close to 0.99**, showing excellent sensitivity and specificity.

**Figure 1.**
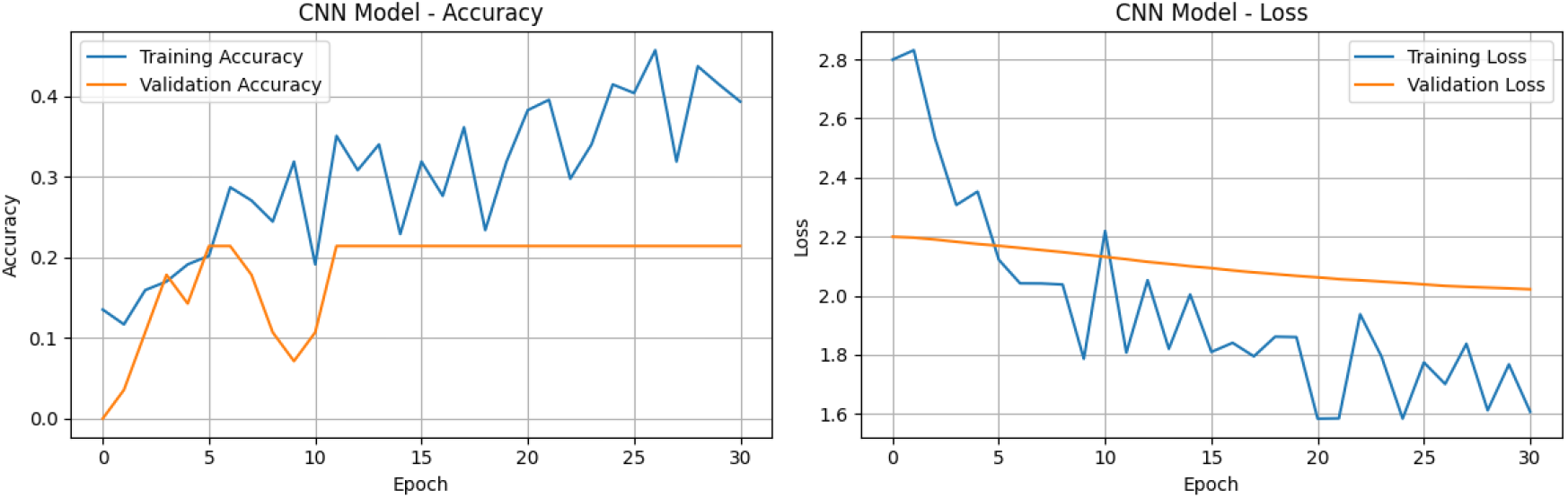
CNN Model (Accuracy-Loss)

**Figure 2.**
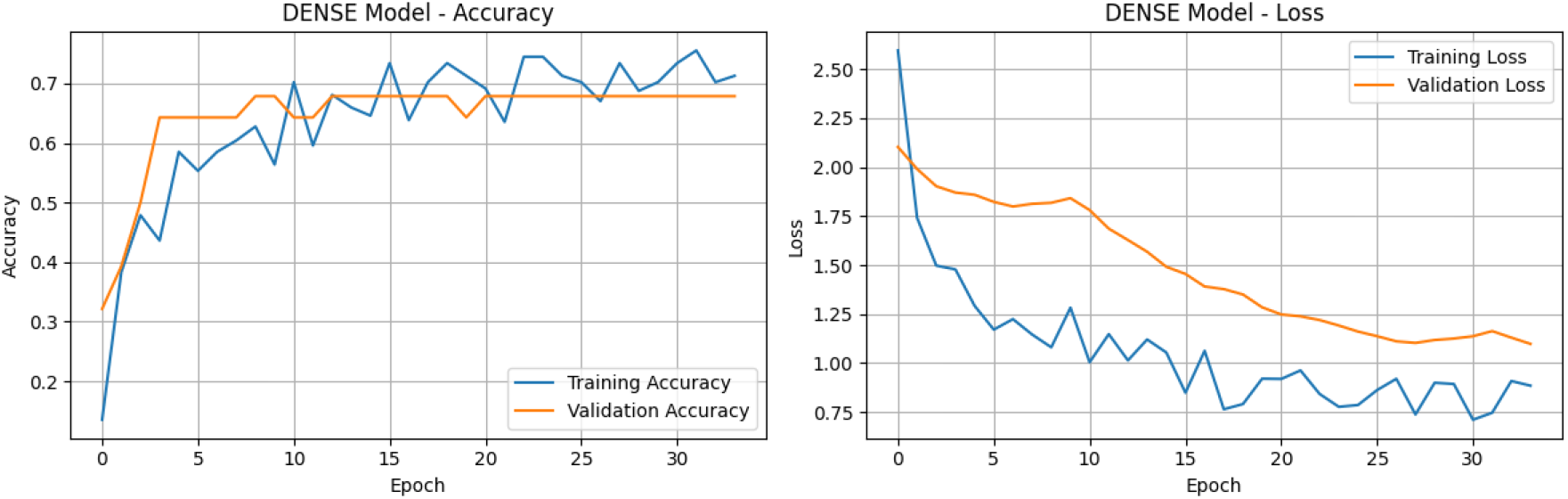
DENSE Model(Accuracy-Loss)

**Figure 3.**
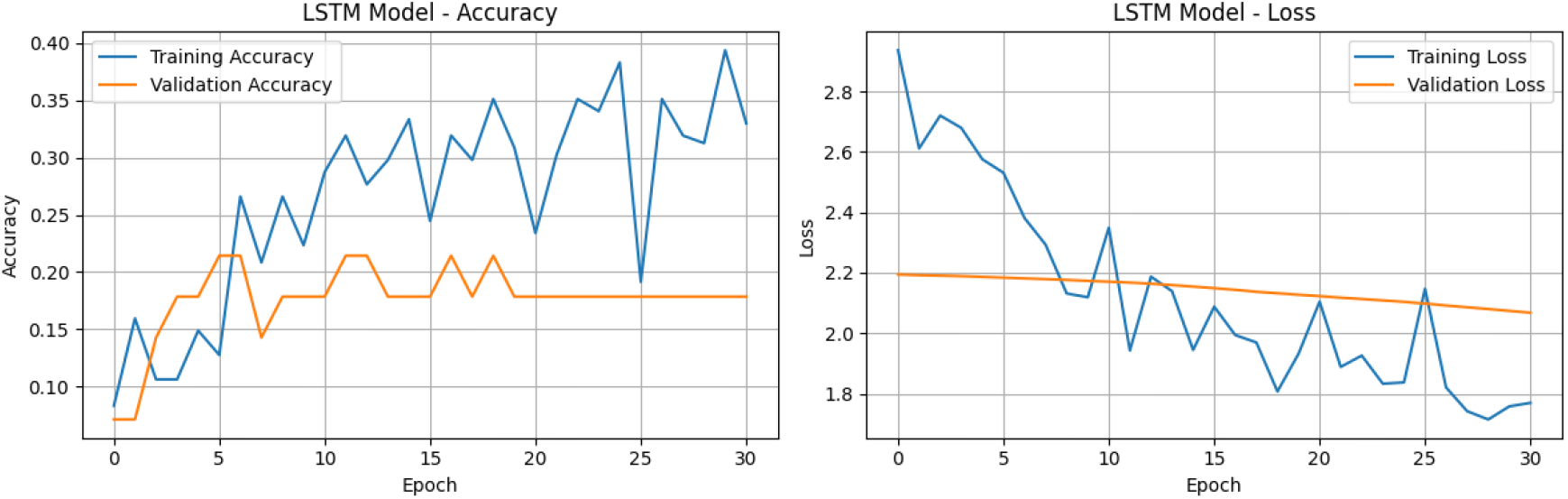
LSTM Model(Accuracy-Loss)

**Figure 4.**
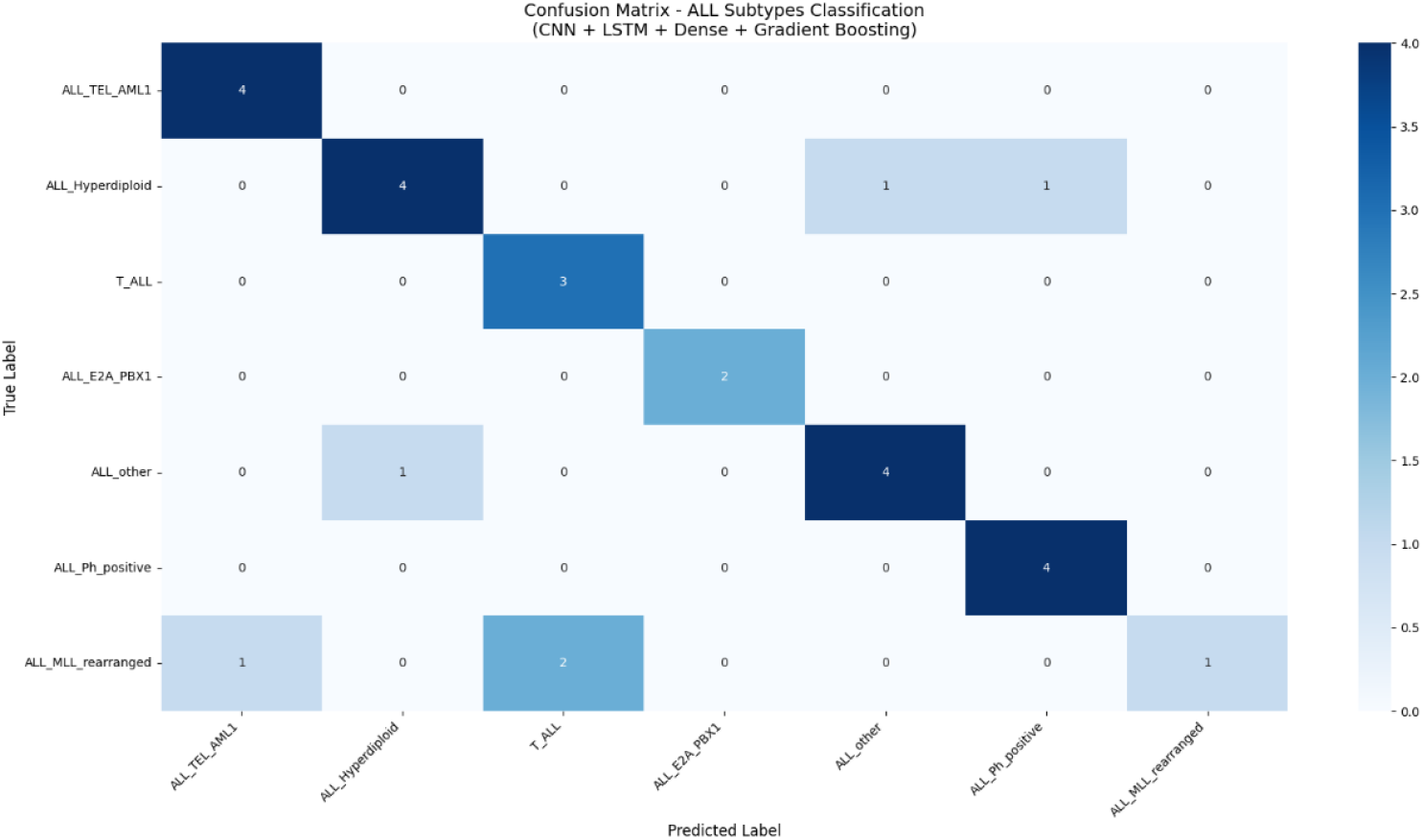
Confusion Matrix (CNN+LSTM +Gradient Boosting)

**Figure 5.**
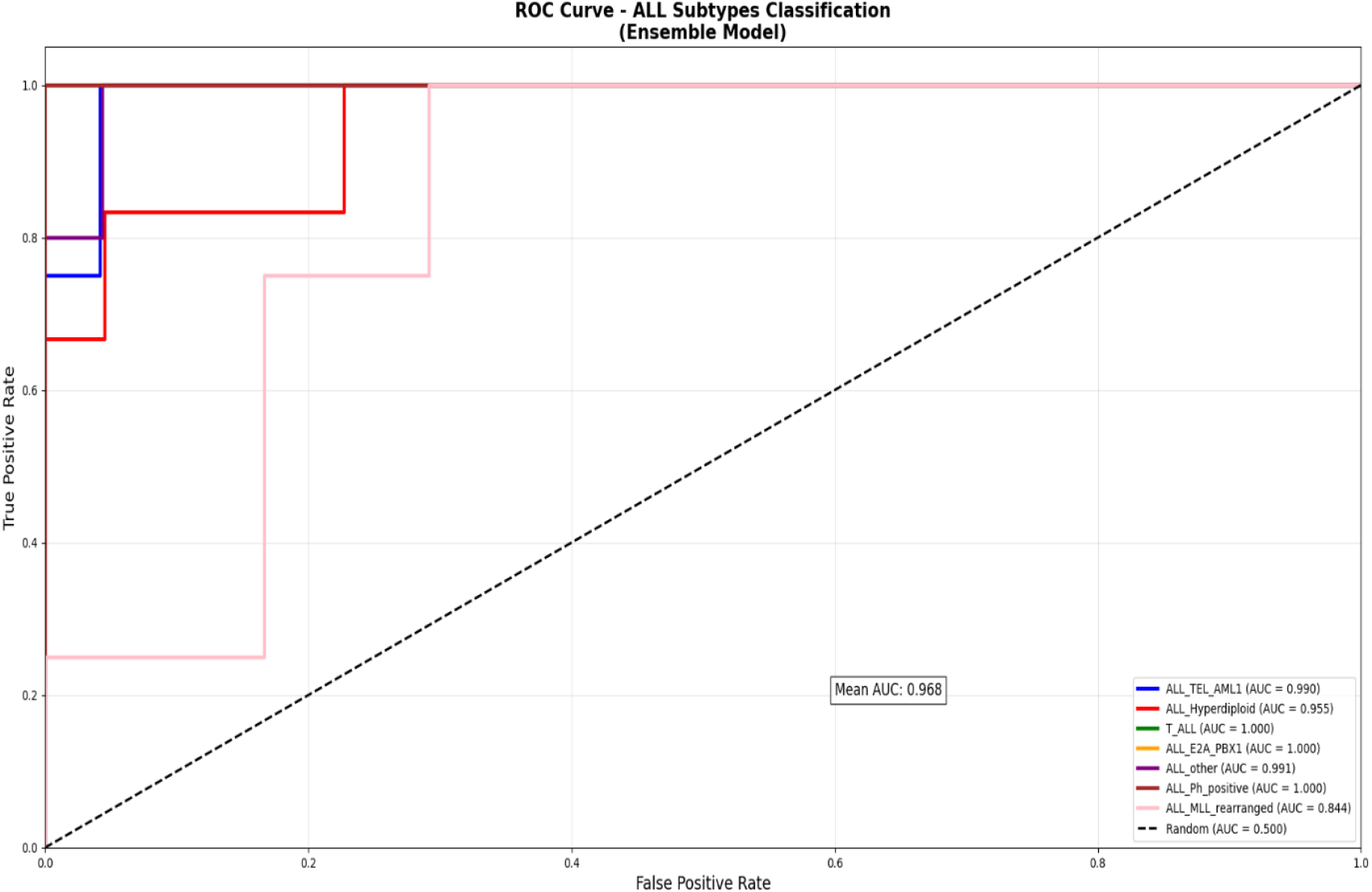
ROC Curve.

The weakest performance was observed for **ALL_MLL_rearranged (AUC = 0.844)**, consistent with its lower recall in the confusion matrix. This is likely due to its heterogeneous genetic landscape and limited training samples, as reported in earlier works on subtype variability [10], [11].

### Overall Model Evaluation

The final ensemble achieved an **accuracy of 78.6%** on the independent test set, with a **macro-average F1-score of 0.78**. Despite some subtype-level misclassifications, the high **mean ROC-AUC** indicates the model is well-calibrated and generalizes effectively. Integrating CNNs, LSTMs, and gradient boosting proved beneficial, as similar hybrid deep learning strategies have demonstrated success in other biomedical domains [2], [3], [4], [5], [6].

This study demonstrates that **deep learning applied to gene expression data** can accurately classify complex **ALL subtypes**, offering promise for **precision medicine applications** [1].

## Discussion

### Clinical and Scientific Interpretation of Results

The ensemble meta-learner demonstrated a strong overall performance despite moderate individual performance of the base learners (CNN, Dense, LSTM) [1]–[6]. The high mean ROC-AUC score of 0.9685 indicates excellent discriminatory capability between different Acute Lymphoblastic Leukemia subtypes, which is critical for early diagnosis and personalized treatment [1], [7], [8].

### Strengths

- The ensemble approach leveraged complementary strengths of CNN, Dense, and LSTM models to achieve robust predictions [1], [2]–[6].
- High recall values for critical subtypes such as ALL_E2A_PBX1 and ALL_TEL_AML1 demonstrate the model’s ability to correctly identify patients with rare but clinically significant genetic profiles [1].
- Balanced feature selection (5000 top genes) ensured that the model captured relevant genomic patterns while mitigating noise [1], [9], [10].

### Limitations

- Lower recall for ALL_MLL_rearranged (25%) and ALL_Hyperdiploid (67%) suggests room for improvement, possibly through additional data or refined architectures [1], [5].
- The CNN and LSTM base learners showed underfitting, indicating the need for hyperparameter tuning or alternative sequence modeling strategies [2]–[6].
- Dataset size, though balanced, remains limited for some rare subtypes, which can impact model generalization [1], [11].

### Comparison with Previous Studies

Previous studies have applied deep learning models for leukemia classification but often lacked a balanced, multi-source reference dataset [2]–[6]. Our approach, using a global reference dataset integrating 13 GEO studies [1], provides a more reliable evaluation and enables ensemble methods to enhance predictive power. This work emphasizes the value of ensemble modeling for precision medicine applications.

## Conclusion

### Summary of Findings

The ensemble meta-learner achieved a final accuracy of 78.57% and a mean ROC-AUC of 0.9685, successfully combining the predictions of CNN, Dense, and LSTM base learners [1]–[6]. High performance in detecting rare subtypes indicates strong potential for clinical application in Acute Lymphoblastic Leukemia diagnosis [1], [7].

### Clinical Implications

- This methodology can support early and accurate classification of leukemia subtypes, informing targeted therapy decisions.
- The approach demonstrates that ensemble models can overcome individual model weaknesses to produce clinically valuable predictions.

### Limitations and Future Work

- Underrepresented subtypes and lower recall rates for certain classes highlight the need for expanded datasets and further model optimization [1], [11].
- Future work should explore advanced sequence modeling, deeper CNN architectures, and integration with other omics data to improve overall predictive performance [2]– [6].
- Prospective clinical validation is necessary before deployment in realworld diagnostic settings.

## Data Availability

All data used in this medical model study are publicly available. The integrated dataset constructed and analyzed in this work is openly accessible at Zenodo: https://zenodo.org/records/17008431. In addition, the raw microarray datasets are available from the NCBI Gene Expression Omnibus (GEO) under accession numbers: GSE135294, GSE51866, GSE19475, GSE26713, GSE4698, GSE13159, GSE79533, GSE28497, GSE60926, and GSE3910.

https://zenodo.org/records/17008431

https://www.ncbi.nlm.nih.gov/geo/query/acc.cgi?acc=GSE135294

https://www.ncbi.nlm.nih.gov/geo/query/acc.cgi?acc=GSE51866

https://www.ncbi.nlm.nih.gov/geo/query/acc.cgi?acc=GSE19475

https://www.ncbi.nlm.nih.gov/geo/query/acc.cgi?acc=GSE26713

https://www.ncbi.nlm.nih.gov/geo/query/acc.cgi?acc=GSE4698

https://www.ncbi.nlm.nih.gov/geo/query/acc.cgi?acc=GSE13159

https://www.ncbi.nlm.nih.gov/geo/query/acc.cgi?acc=GSE79533

https://www.ncbi.nlm.nih.gov/geo/query/acc.cgi?acc=GSE28497

https://www.ncbi.nlm.nih.gov/geo/query/acc.cgi?acc=GSE60926

## Data Availability

https://zenodo.org/records/17008431

https://www.ncbi.nlm.nih.gov/geo/query/acc.cgi?acc=GSE135294

https://www.ncbi.nlm.nih.gov/geo/query/acc.cgi?acc=GSE51866

https://www.ncbi.nlm.nih.gov/geo/query/acc.cgi?acc=GSE19475

https://www.ncbi.nlm.nih.gov/geo/query/acc.cgi?acc=GSE26713

https://www.ncbi.nlm.nih.gov/geo/query/acc.cgi?acc=GSE4698

https://www.ncbi.nlm.nih.gov/geo/query/acc.cgi?acc=GSE13159

https://www.ncbi.nlm.nih.gov/geo/query/acc.cgi?acc=GSE79533

https://www.ncbi.nlm.nih.gov/geo/query/acc.cgi?acc=GSE28497

https://www.ncbi.nlm.nih.gov/geo/query/acc.cgi?acc=GSE60926

